# Short-term and long-term outcome prediction for patients with coronary artery disease using machine learning and comprehensive multi-center patient data

**DOI:** 10.1101/2025.05.26.25328366

**Authors:** Emma Bogner, Bryan Har, Bing Li, Danielle A. Southern, Christopher L. F. Sun, Robert C. Welsh, Benjamin Tyrrell, Colm J. Murphy, Arjun Puri, Joon Lee

**Author notes:** Corresponding author: Joon Lee, PhD, Cal Wenzel Precision Health Building Room 5E17, 3280 Hospital Dr NW, Calgary, Alberta, Canada T2N 4Z6. Conflicts of interest: AP and JL are the co-founders and major shareholders of Symbiotic AI, Inc. BH and CJM are minor shareholders of Symbiotic AI, Inc.

## Abstract

**Background:** Revascularization decision-making for patients with coronary artery disease (CAD) can benefit from accurate patient outcome prediction. While previous studies have employed data-driven methods including machine learning (ML) to develop prediction models, they were mostly based on small patient cohorts with strict inclusion and exclusion criteria, limited feature sets, and only internal validation.

**Objectives:** To develop and externally validate ML-based models to predict a wide range of short- and long-term outcomes for patients with obstructive CAD using large-scale multi-center patient data.

**Methods:** Comprehensive data from patients with obstructive CAD who underwent coronary angiography at three hospitals in Alberta, Canada between 2009 and 2019 were extracted from the APPROACH Registry and linked administrative health databases. To predict all-cause mortality and major adverse cardiovascular events at 90 days, 1 year, 3 years, and 5 years, over 12,000 features were considered in an extensive ML framework that employed rigorous hyperparameter tuning, calibration, algorithmic bias assessment, and external validation. In addition to traditional ML models, we employed a generative transformer-based tabular foundation model, TabPFN. To increase the clinical utility of these prediction models, we also performed a secondary analysis that investigated the impact of the exclusion of angiography data on prediction performance.

**Results:** A total of 44,462 catheterizations from 38,767 unique patients were included in the study. The median areas under the receiver operating characteristic curves of the best models, mostly TabPFNs, in external validation ranged from 0.797 to 0.845 and 0.694 to 0.753 for mortality and MACE, respectively. CAD factors, angiography results, and patient history were the most influential feature groups. The algorithmic bias assessment focusing on patient sex showed that the models were mostly fair. The secondary analysis showed that prediction performance degraded slightly when angiography features were excluded.

**Conclusions:** The prediction performance reported in this study is state-of-the-art compared to previous studies. The large sample size, extensive feature set, external validation, and transformer architecture led to personalized models with robust performance. The models from this study have the potential to improve coronary revascularization decision-making and patient outcomes via accurate prognosis.

**Condensed Abstract:** We developed and externally validated machine learning models to predict short- and long-term outcomes in patients with obstructive coronary artery disease using comprehensive data from over 44,000 catheterizations from over 38,000 patients across three Canadian hospitals. Using hundreds of features and advanced models including a transformer-based tabular foundation model, we achieved state-of-the-art performance in predicting mortality and major adverse cardiovascular events. Key predictors included CAD factors, angiography results, and patient history. The models showed minimal algorithmic bias by sex and retained good accuracy even without angiography data. These results suggest significant potential to enhance clinical decision-making and patient prognosis.

## Introduction

Treatment decision-making for patients with coronary artery disease (CAD) can be challenging particularly in the presence of complex multivessel anatomy or extensive comorbidities. Although a number of randomized controlled trials (RCTs) have been conducted to guide coronary revascularization decision-making,^1^ unique individual patient characteristics often make it difficult to apply the existing clinical practice guidelines that are optimized for typical patients. While risk scores such as SYNTAX II, which has recently been upgraded to the SYNTAX score II 2020,^2^ can provide decision support via quantification of the severity of CAD, their predictive performances remain limited.

Recognizing that it is impractical to conduct an RCT for every unique CAD phenotype, machine learning (ML) presents an effective, data-driven method to learn from large-scale patient data and operationalize the learned knowledge in the form of a prediction model. Such a model can ingest a new patient’s data and generate personalized decision support insights. Several recent studies utilized ML to predict a variety of short- and long-term mortality and major adverse cardiovascular event (MACE) outcomes for various patient cohorts with CAD, reporting prediction performances surpassing those of SYNTAX II or the SYNTAX score II 2020.^3–6^ However, their predicted outcomes were often too short-term to meaningfully support treatment decisions (e.g., 30-day mortality^4^ or 6-month MACE^3^). Also, some of these studies used a rather small pool of predictor variables (e.g., 21^3^ or 30^5^).

We aimed to develop truly personalized mortality and MACE prediction models based on hundreds of features (i.e., predictor variables) from tens of thousands of patients with CAD from three hospitals in Alberta, Canada. Unlike the previous ML CAD studies, we focused on constructing and utilizing rich patient history information from the 5-year period prior to the index coronary angiography, in addition to typical demographic and CAD-related clinical data. Our predicted outcomes ranged from 90-day to 5-year endpoints, and prediction performance was validated with one of the three sites serving as an external site.

## Methods

### Data Sources

This study utilized retrospective data from three hospitals in Alberta, Canada. The primary data source used in this study was the Alberta Provincial Project for Outcome Assessment in Coronary Heart disease (APPROACH) registry,^7^ which captures demographic and clinical variables (e.g., comorbidities, referral indications, pre- and peri-procedural details) for all patients undergoing diagnostic cardiac catheterization in Alberta. Standardized diagnostic angiography results, including measures of the location and extent of coronary lesions, were derived from CARAT (a specialized CAD annotation and reporting software) data in the form of the 17-segment heart model and linked to the corresponding patient records.

International Classification of Diseases, Canadian 10^th^ Edition (ICD-10-CA) diagnosis codes and Canadian Classification of Health Interventions (CCI) procedure codes were extracted from administrative databases capturing hospital admissions (Discharge Abstract Database)^8^ and ambulatory care visits (National Ambulatory Care Reporting System)^9^ and used to define patient history and identify outcome events. The dates of death recorded in the Vital Statistics database were used to identify out-of-hospital mortality events.

### Patient Cohort

Adult patients (≥18 years) were considered eligible for inclusion in our study if they underwent a cardiac catheterization at one of the three cardiac catheterization sites in Alberta between January 2009 and March 2019 and with a definitive obstructive CAD diagnosis, defined as any stenosis ≥50% in the left main coronary artery and/or ≥70% in any other coronary artery. Patients were excluded if their primary residence was outside of Alberta or if the follow-up period for their index catheterization fell outside of the data collection window to ensure complete outcome data. Patients presenting with ST-segment elevation myocardial infarction (STEMI) were also excluded due to the emergent nature of these cases, where the clinical guidelines for treating culprit lesions with immediate PCI are already well established.

### Predicted Patient Outcomes

We targeted two outcomes, all-cause mortality and MACE, at four timepoints, including 90 days, 1 year, 3 years, and 5 years from treatment delivery, for a total of eight predicted outcomes. Follow-up start was specified as the procedure date for patients who underwent a revascularization intervention (percutaneous coronary intervention [PCI] or coronary artery bypass grafting [CABG]) and the index catheterization date for those who received medical therapy only. MACE was defined as a five-point composite including acute myocardial infarction, heart failure, stroke, repeat revascularization, and all-cause mortality (to address competing risks). The individual MACE endpoints were defined via the identification of ICD-10-CA diagnosis codes and CCI procedure codes in administrative data.

### Model Development and Validation

We defined three modelling cohorts using different data partitioning schemas based on site to allow for external validation of our models. In each cohort, data from one site was designated as the testing set, while data from the other two sites were pooled and split again into training (85%) and calibration (15%) sets. Partitioning was carried out at the patient level to avoid distributing procedures from the same patient between sets. All splits were stratified by prediction target to ensure consistent event rates across sets.

A set of 200 features (200 chosen based on preliminary analysis that experimented different numbers of features) were selected for each model, using *SelectFromModel* in in the *scikit-learn* Python package with random forest as the base model, from a pool of 12,112 derived and engineered features comprising patient demographics, modifiable lifestyle factors (e.g., smoking status, body mass index [BMI]), comorbidities, angiography results, CAD factors (e.g., index catheterization indication, procedure priority, prior revascularizations, family history of CAD, Canadian Cardiovascular Society angina score), and 5-year diagnostic and procedural histories relative to the patient’s index coronary angiography. Features missing more than 25% of the time were excluded from selection. All continuous and categorical features were linearly normalized or one-hot encoded, respectively. Missing values were imputed using the mean of the training cohort for continuous features or the mode of the training cohort for categorical features.

As a secondary analysis, we also trained models without angiography data. There are two motivations: 1) without angiography data, our models can be used for a larger patient population more upstream in the CAD patient care flow prior to coronary angiography; and 2) unless automated angiogram image analysis is used for stenosis assessment,^10,11^ which is not currently used in real-world patient care, angiography results would have to be entered manually before predictions can be generated.

We trained six models for each cohort/outcome combination: logistic regression, random forest, extreme gradient boosting, deep feed-forward neural network, the base Tabular Prior-data Fitted Network^12^ (TabPFN), and TabPFN’s subsampled ensemble variant. TabPFN is a generative transformer-based foundation model for tabular data pre-trained on 100 million synthetic data sets. TabPFN leverages the same in-context learning used by large language models to perform training and inference for any tabular dataset in a single pass, without any need for hyperparameter tuning. The base model is the main TabPFN model, whereas the subsampled ensemble variant incorporates multiple TabPFN models into an ensemble, with each receiving a different subset of the training data. For both types of TabPFN, we utilized TabPFN’s built-in preprocessing, imputation, and class imbalance mitigation functionalities.

For all models except TabPFN, the best hyperparameters and architectures were selected for each algorithm based on the results of 10-fold cross-validation within training data by taking the model with the highest median area under the precision–recall curve (AUPRC) to account for low event rates (see Table A in the Appendix for the hyperparameter search spaces). All models, including TabPFN, were then calibrated on the held-out calibration set using isotonic regression. The calibration set was also used to set the decision threshold for each model as required to maximize F1 scores. The performance of all calibrated models was then externally validated with respect to accuracy, area under the receiver operating characteristic curve (AUROC), AUPRC, positive predictive value (PPV), negative predictive value (NPV), sensitivity, specificity, and Brier score at their respective testing sites using a bootstrapped analysis to generate median values with 95% confidence intervals. A final best performing model for each outcome-cohort combination was selected by choosing the algorithm with the highest median AUROC and AUPRC during external validation.

For all models except TabPFN, to mitigate class imbalance, the minority class was up-sampled using the synthetic minority oversampling technique (SMOTE)^13^ to match the size of the majority class for outcomes with event rates less than 40%.

Furthermore, we used SHapley Additive exPlanations (SHAP)^14^ to quantify the importance of the main feature categories included in the final models by summing the absolute mean SHAP values of the individual features and calculating percentage contribution by category.

Finally, we used the Aequitas toolkit^15^ to assess the fairness of the best models with respect to patient sex (patient ethnicity or race was unavailable in our data set). Disparities between male and female patients were assessed in terms of false positive rate (FPR), true positive rate (TPR), and PPV, with male being the reference group. The default threshold of 0.8 in Aequitas was used to make dichotomous fairness decisions.

All ML modeling was conducted in Python using scikit-learn, PyTorch, XGBoost, imbalanced-learn, Optuna, shap, shapiq, and tabpfn.

### Ethics Approval

The study was approved by the Conjoint Health Research Ethics Board at the University of Calgary (REB20-1879) and carried out in accordance with the TRIPOD+AI guidelines (see the Appendix). Informed consent was waived due to the retrospective nature of data collection and the impracticality of obtaining consent from the large number of patients involved.

## Results

A total of 60,335 diagnostic cardiac catheterizations from 51,506 unique patients, excluding those with an indication of STEMI, were recorded in the APPROACH database between 2009 and 2019. Of these, 44,462 catheterizations from 38,767 unique patients met the eligibility criteria for this study. The final cohort sizes across the three sites for each outcome varied due to different follow-up periods and are shown in Figure 1.

**Figure 1.**
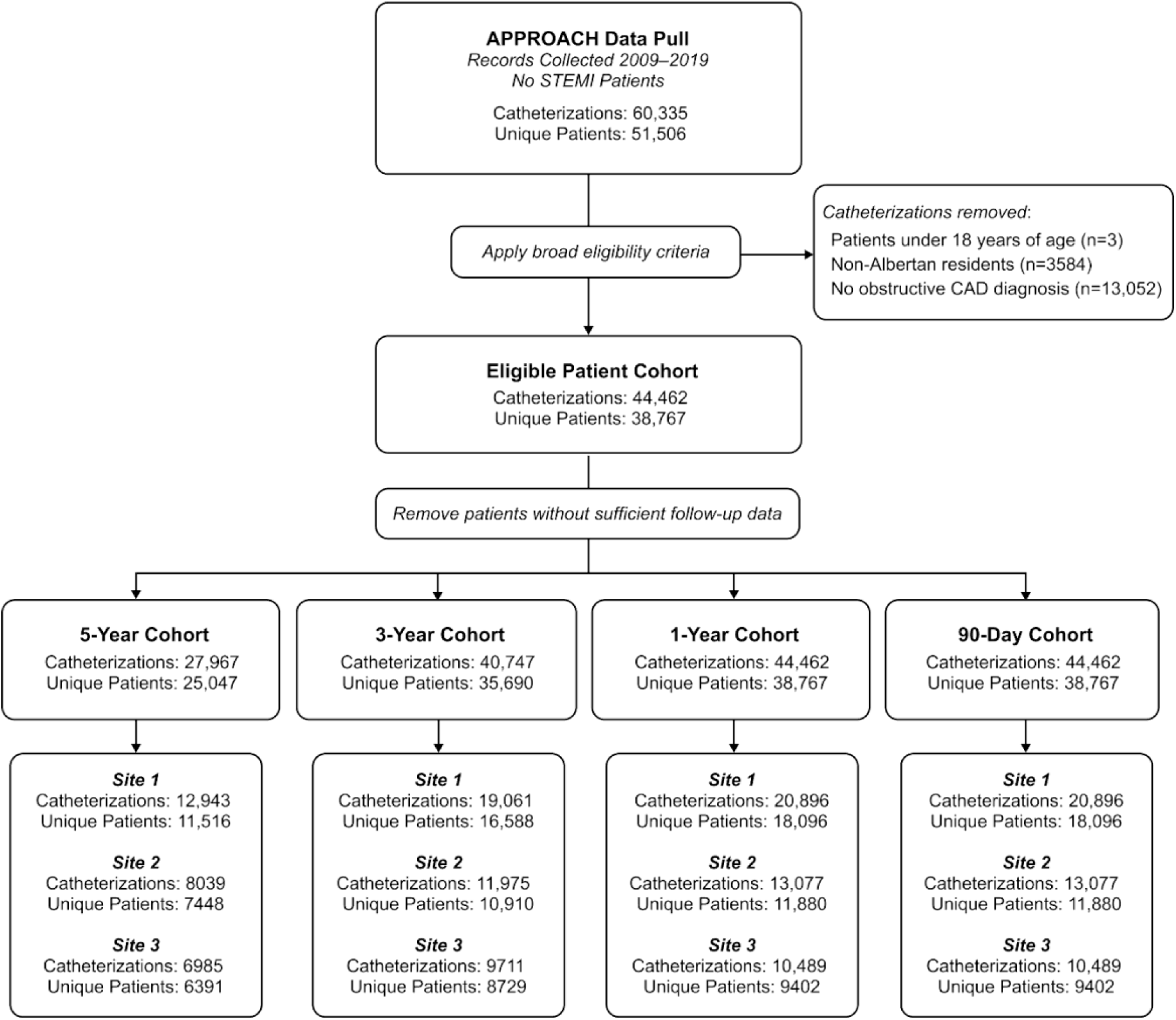
Patient cohort flow chart.

Table 1 describes the baseline patient characteristics of the eligible cohort, stratified by site. The majority of eligible index catheterizations were performed at Site 1 (47%), followed by Site 2 (29.4%) and Site 3 (23.6%). Across all sites, the median age of eligible patients was approximately 66 years, where most patients were between the ages of 58 and 75.

**Table 1.** Eligible patient cohort (44,462 catheterizations) characteristics per index catheterization, stratified by catheterization site.

| **Variable** | **Site 1**<br>**(N = 20,896)** | **Site 2**<br>**(N = 13,077)** | **Site 3**<br>**(N = 10,489)** |
| --- | --- | --- | --- |
| Age, years, median (IQR) | 67.0 (59.0–74.9) | 66.2 (58.0–74.7) | 66.0 (57.9–74.5) |
| Sex, *n* (%) |  |  |  |
| Male | 16,162 (77.3) | 9948 (76.1) | 7923 (75.5) |
| Female | 4734 (22.7) | 3129 (23.9) | 2566 (24.5) |
| Indication, *n* (%) |  |  |  |
| NSTEMI | 8687 (41.6) | 6638 (50.8) | 5690 (54.2) |
| Unstable Angina | 5291 (25.3) | 2105 (16.1) | 2143 (20.4) |
| Stable Angina | 4086 (19.6) | 3276 (25.1) | 1300 (12.5) |
| Other | 2704 (12.9) | 1043 (7.9) | 1312 (12.5) |
| Unknown | 128 (0.6) | 15 (0.1) | 44 (0.4) |
| Coronary Artery Disease Severity, *n* (%) |  |  |  |
| Single Vessel | 4221 (20.3) | 3077 (23.5) | 1790 (17.1) |
| Double Vessel | 5548 (26.5) | 3724 (28.5) | 2637 (25.1) |
| Triple Vessel | 8065 (38.6) | 4560 (34.9) | 4398 (41.9) |
| Left Main | 3062 (14.6) | 1716 (13.1) | 1664 (15.9) |
| Revascularization Intervention, *n* (%) |  |  |  |
| PCI | 10,728 (51.7) | 8783 (67.3) | 6234 (59.7) |
| CABG | 3482 (16.8) | 1812 (13.9) | 1812 (17.3) |
| Medical Therapy Only | 6544 (31.5) | 2464 (18.9) | 2401 (23.0) |
| Comorbidities, *n* (%) |  |  |  |
| Diabetes | 6606 (31.6) | 3399 (26.0) | 3322 (31.7) |
| Hypertension | 14,615 (69.9) | 7244 (55.4) | 7401 (70.6) |
| Chronic Lung Disease | 3060 (14.6) | 1105 (8.4) | 622 (5.9) |
| Heart Failure | 1728 (8.3) | 812 (6.2) | 827 (7.9) |
| CKD | 884 (4.23) | 105 (0.8) | 173 (1.6) |
| Dyslipidemia | 14,961 (71.6) | 7164 (54.8) | 8001 (76.3) |
| Atrial Fibrillation | 827 (4.0) | 116 (0.8) | 79 (0.8) |
| CEVD | 1050 (5.0) | 508 (3.9) | 440 (4.2) |
| Smoking History, *n* (%) |  |  |  |
| Never Smoked | 9796 (46.9) | 7163 (54.7) | 4468 (42.6) |
| Past Smoker | 6773 (32.4) | 3187 (24.4) | 3363 (32.1) |
| Current Smoker | 4327 (20.7) | 2727 (20.9) | 2658 (25.3) |
| Prior PCI Procedures, *n* (%) |  |  |  |
| None | 15,673 (75.0) | 9928 (75.9) | 7875 (75.1) |
| One | 3418 (16.4) | 1991 (15.3) | 1712 (16.3) |
| Multiple | 1805 (8.60) | 1158 (8.80) | 902 (8.60) |
| Prior CABG Procedures, *n* (%) |  |  |  |
| None | 18,882 (90.4) | 11,914 (91.1) | 9311 (88.8) |
| One | 1894 (9.1) | 1120 (8.6) | 1126 (10.7) |
| Multiple | 120 (0.50) | 43 (0.30) | 52 (0.50) |
| Prior Myocardial Infarctions, *n* (%) |  |  |  |
| None | 19,830 (94.9) | 12,261 (93.8) | 9588 (91.4) |
| One | 907 (4.40) | 736 (5.60) | 823 (7.90) |
| Multiple | 159 (0.70) | 80 (0.60) | 78 (0.70) |
| Event Rates, *n* (%) |  |  |  |
| 5-Year MACE | 8338 (39.9) | 5701 (43.6) | 4447 (42.4) |
| 3-Year MACE | 6665 (31.9) | 4485 (34.3) | 3682 (35.1) |
| 1-Year MACE | 4242 (20.3) | 3047 (23.3) | 2591 (24.7) |
| 90-Day MACE | 1964 (9.4) | 1739 (13.3) | 1521 (14.5) |
| 5-Year Mortality | 3051 (14.6) | 2105 (16.1) | 1689 (16.1) |
| 3-Year Mortality | 1985 (9.5) | 1334 (10.2) | 1196 (11.4) |
| 1-Year Mortality | 899 (4.3) | 667 (5.1) | 619 (5.9) |
| 90-Day Mortality | 460 (2.2) | 314 (2.4) | 294 (2.8) |

CABG—coronary artery bypass graft; CEVD—cerebrovascular disease; CKD—chronic kidney disease; IQR—interquartile range; MACE—major adverse cardiovascular events; NSTEMI—non-ST-segment elevation myocardial infarction; PCI—percutaneous coronary intervention.

Most patients had no prior history of acute myocardial infarction or related revascularization procedures, presented with NSTEMI or unstable angina, and received PCI as an index intervention. Hypertension, dyslipidemia, and diabetes were commonly reported as comorbidities in all patients. However, the overall prevalence of all comorbidities was much lower in patients from Site 2. In addition, Sites 2 and 3 were found to have slightly higher event rates than Site 1 for all outcomes, with the greatest disparity observed between MACE outcomes.

Table 2 tabulates the prediction performance of the best model for each outcome and test cohort. All mortality models performed well in external validation, achieving AUROC scores between 0.797 and 0.845, and AUPRC scores between 0.054 and 0.344. The MACE models showed more moderate performance, with lower AUROCs (0.694–0.753) but higher AUPRCs (0.146–0.561) overall. For most outcome-test cohort combinations (21 out of 24), the best model was a TabPFN. For both MACE and mortality, model performance tended to decrease moving from 5-year to 90-day outcomes in terms of AUPRC, PPV, and TPR, as expected from decreasing events rates.

**Table 2.** Prediction performances of best models selected in external validation. All metrics are provided as median values with 95% confidence intervals.

| Test Cohort | Best Model | AUROC | AUPRC | Accuracy | PPV | NPV | TPR | TNR |
| --- | --- | --- | --- | --- | --- | --- | --- | --- |
| **5-Year All-Cause Mortality** |  |  |  |  |  |  |  |  |
| 1 | TabPFN (subsample) | 0.813 (0.808, 0.821) | 0.295 (0.279, 0.301) | 0.774 (0.766, 0.778) | 0.364 (0.344, 0.370) | 0.935 (0.930, 0.939) | 0.684 (0.667, 0.696) | 0.790 (0.780, 0.794) |
| 2 | TabPFN (base) | 0.822 (0.814, 0.832) | 0.333 (0.313, 0.344) | 0.818 (0.814, 0.830) | 0.461 (0.434, 0.473) | 0.912 (0.909, 0.920) | 0.575 (0.561, 0.583) | 0.868 (0.862, 0.875) |
| 3 | TabPFN (subsample) | 0.845 (0.831, 0.846) | 0.344 (0.320, 0.359) | 0.823 (0.816, 0.827) | 0.467 (0.439, 0.481) | 0.918 (0.915, 0.920) | 0.593 (0.575, 0.616) | 0.868 (0.861, 0.872) |
| **3-Year All-Cause Mortality** |  |  |  |  |  |  |  |  |
| 1 | TabPFN (base) | 0.796 (0.791, 0.807) | 0.198 (0.189, 0.205) | 0.797 (0.793, 0.801) | 0.262 (0.252, 0.270) | 0.952 (0.950, 0.955) | 0.614 (0.600, 0.632) | 0.815 (0.813, 0.821) |
| 2 | TabPFN (base) | 0.802 (0.795, 0.810) | 0.211 (0.194, 0.223) | 0.820 (0.815, 0.827) | 0.301 (0.280, 0.314) | 0.942 (0.939, 0.947) | 0.546 (0.525, 0.572) | 0.852 (0.845, 0.857) |
| 3 | TabPFN (base) | 0.817 (0.801, 0.827) | 0.254 (0.229, 0.262) | 0.858 (0.857, 0.871) | 0.414 (0.385, 0.428) | 0.926 (0.925, 0.934) | 0.456 (0.423, 0.475) | 0.912 (0.912, 0.921) |
| **1-Year All-Cause Mortality** |  |  |  |  |  |  |  |  |
| 1 | TabPFN (base) | 0.797 (0.791, 0.803) | 0.097 (0.093, 0.100) | 0.884 (0.879, 0.884) | 0.165 (0.161, 0.172) | 0.973 (0.972, 0.973) | 0.431 (0.423, 0.447) | 0.904 (0.899, 0.904) |
| 2 | TabPFN (subsample) | 0.810 (0.798, 0.812) | 0.122 (0.116, 0.126) | 0.895 (0.890, 0.898) | 0.219 (0.214, 0.229) | 0.966 (0.966, 0.969) | 0.416 (0.400, 0.426) | 0.922 (0.916, 0.922) |
| 3 | TabPFN (subsample) | 0.830 (0.809, 0.835) | 0.137 (0.125, 0.149) | 0.796 (0.790, 0.802) | 0.175 (0.160, 0.188) | 0.976 (0.974, 0.979) | 0.684 (0.652, 0.708) | 0.802 (0.798, 0.808) |
| **90-Day All-Cause Mortality** |  |  |  |  |  |  |  |  |
| 1 | TabPFN (base) | 0.821 (0.803, 0.829) | 0.054 (0.046, 0.066) | 0.960 (0.956, 0.961) | 0.165 (0.144, 0.195) | 0.983 (0.981, 0.984) | 0.223 (0.196, 0.261) | 0.976 (0.973, 0.978) |
| 2 | TabPFN (base) | 0.810 (0.801, 0.821) | 0.060 (0.056, 0.067) | 0.926 (0.923, 0.929) | 0.130 (0.119, 0.136) | 0.984 (0.982, 0.984) | 0.370 (0.332, 0.381) | 0.940 (0.936, 0.942) |
| 3 | TabPFN<br>(subsample) | 0.838 (0.816, 0.847) | 0.073 (0.064, 0.083) | 0.886 (0.880, 0.891) | 0.119 (0.109, 0.132) | 0.985 (0.983, 0.986) | 0.502 (0.448, 0.531) | 0.897 (0.891, 0.901) |
| **5-Year MACE** |  |  |  |  |  |  |  |  |
| 1 | TabPFN<br>(subsample) | 0.746 (0.739, 0.753) | 0.537 (0.529, 0.549) | 0.697 (0.690, 0.702) | 0.621 (0.611, 0.630) | 0.746 (0.742, 0.756) | 0.621 (0.617, 0.636) | 0.743 (0.738, 0.750) |
| 2 | TabPFN<br>(subsample) | 0.734 (0.728, 0.741) | 0.561 (0.551, 0.577) | 0.684 (0.677, 0.688) | 0.657 (0.645, 0.673) | 0.700 (0.694, 0.706) | 0.570 (0.558, 0.583) | 0.772 (0.763, 0.779) |
| 3 | TabPFN<br>(subsample) | 0.753 (0.739, 0.766) | 0.557 (0.541, 0.573) | 0.690 (0.677, 0.704) | 0.629 (0.611, 0.648) | 0.737 (0.725, 0.753) | 0.647 (0.635, 0.664) | 0.722 (0.705, 0.733) |
| **3-Year MACE** |  |  |  |  |  |  |  |  |
| 1 | TabPFN<br>(subsample) | 0.730 (0.725, 0.737) | 0.437 (0.426, 0.448) | 0.685 (0.681, 0.691) | 0.506 (0.492, 0.516) | 0.806 (0.801, 0.814) | 0.637 (0.630, 0.647) | 0.709 (0.703, 0.712) |
| 2 | XGBoost | 0.734 (0.725, 0.743) | 0.460 (0.451, 0.473) | 0.681 (0.671, 0.688) | 0.526 (0.515, 0.539) | 0.791 (0.779, 0.803) | 0.644 (0.627, 0.659) | 0.699 (0.690, 0.711) |
| 3 | TabPFN<br>(base) | 0.745 (0.737, 0.752) | 0.482 (0.478, 0.491) | 0.696 (0.691, 0.702) | 0.558 (0.552, 0.568) | 0.790 (0.781, 0.796) | 0.640 (0.622, 0.652) | 0.728 (0.718, 0.734) |
| **1-Year MACE** |  |  |  |  |  |  |  |  |
| 1 | Random<br>Forest | 0.707 (0.700, 0.715) | 0.295 (0.286, 0.306) | 0.724 (0.718, 0.729) | 0.374 (0.363, 0.386) | 0.868 (0.862, 0.873) | 0.539 (0.522, 0.554) | 0.771 (0.766, 0.775) |
| 2 | TabPFN<br>(subsample) | 0.720 (0.716, 0.724) | 0.343 (0.341, 0.358) | 0.756 (0.752, 0.758) | 0.474 (0.467, 0.486) | 0.839 (0.836, 0.842) | 0.459 (0.454, 0.484) | 0.841 (0.840, 0.849) |
| 3 | XGBoost | 0.747 (0.734, 0.758) | 0.456 (0.443, 0.473) | 0.644 (0.633, 0.657) | 0.496 (0.481, 0.512) | 0.805 (0.794, 0.816) | 0.732 (0.718, 0.750) | 0.597 (0.584, 0.610) |
| **90-Day MACE** |  |  |  |  |  |  |  |  |
| 1 | TabPFN<br>(subsample) | 0.694 (0.680, 0.696) | 0.146 (0.142, 0.152) | 0.779 (0.778, 0.780) | 0.204 (0.199, 0.212) | 0.939 (0.935, 0.940) | 0.485 (0.463, 0.492) | 0.809 (0.808, 0.811) |
| 2 | TabPFN<br>(base) | 0.724 (0.713, 0.731) | 0.202 (0.194, 0.208) | 0.730 (0.727, 0.736) | 0.259 (0.248, 0.264) | 0.919 (0.916, 0.920) | 0.554 (0.540, 0.570) | 0.755 (0.754, 0.763) |
| 3 | TabPFN<br>(subsample) | 0.707 (0.702, 0.709) | 0.220 (0.210, 0.222) | 0.742 (0.735, 0.745) | 0.290 (0.275, 0.293) | 0.901 (0.900, 0.905) | 0.504 (0.501, 0.514) | 0.784 (0.775, 0.788) |

AUPRC—area under the precision–recall curve; AUROC—area under the receiver operating characteristic curve; MACE—major adverse cardiovascular events; NPV—negative predictive value; PPV—positive predictive value; TPR—true positive rate; TNR—true negative rate.

Table B in the Appendix tabulates the prediction performance results from the secondary analysis that excluded angiography data from the feature pool. Overall, compared to the primary results with angiography data (Table 2), prediction performance decreased slightly in terms of median AUROCs and AUPRCs, although in many cases the 95% confidence intervals overlapped. The majority of the best models (16 out of 24) were again TabPFNs.

Figure 2 shows the calibration plots of each best model. Following isotonic regression, the mortality models showed excellent calibration with Brier scores between 0.02 and 0.10. The Brier scores of the MACE models were slightly higher (0.09–0.21) but still showed reasonable calibration. For both MACE and mortality, the models predicting short-term outcomes were found to be better calibrated than those predicting long-term outcomes.

**Figure 2.**
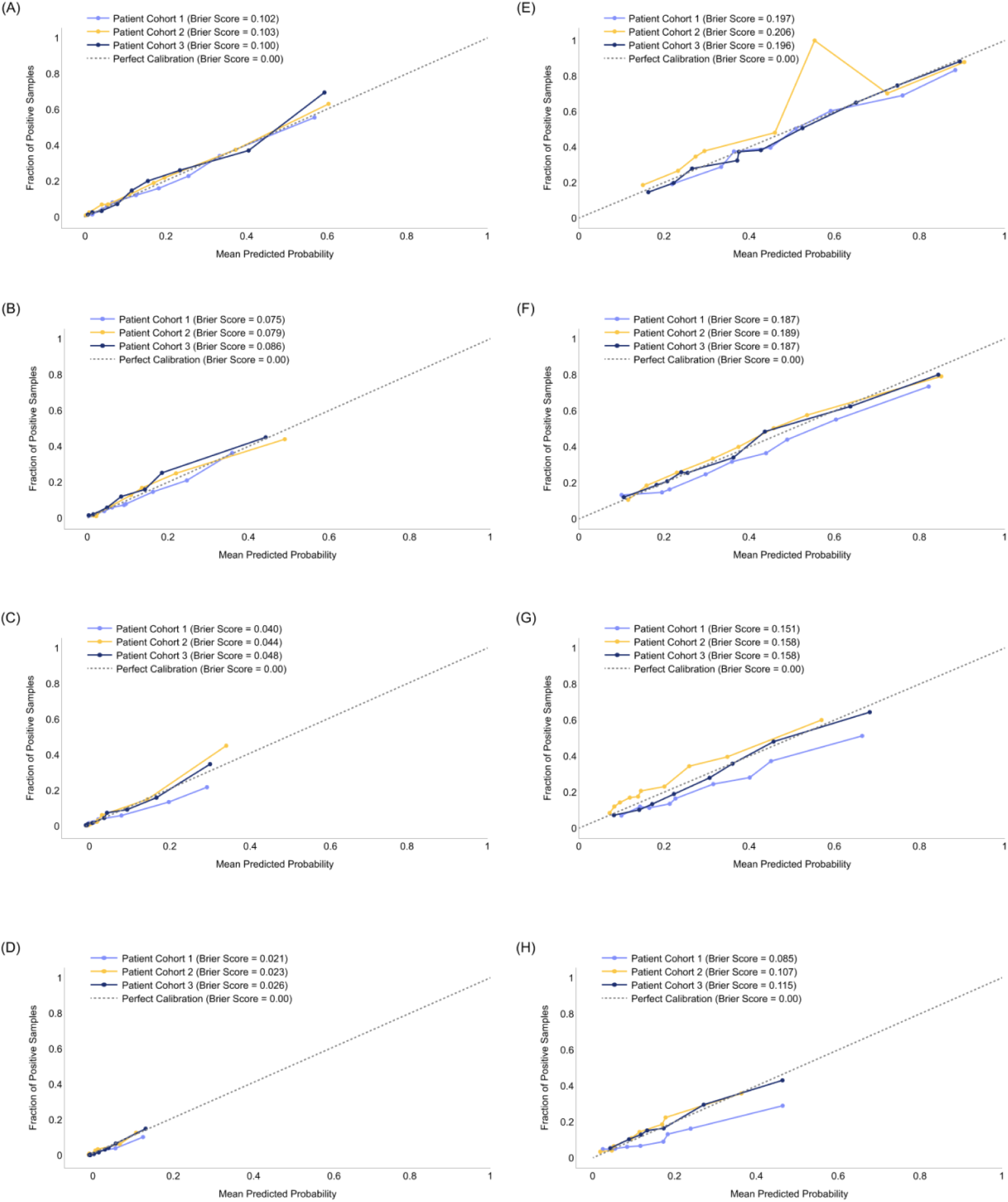
Calibration curves and Brier scores of the best models selected in external validation predicting: (A) 5-year mortality, (B) 3-year mortality, (C) 1-year mortality, (D) 90-day mortality, (E) 5-year MACE, (F) 3-year MACE, (G) 1-year MACE, (H) 90-day MACE.

Figure 3 illustrates the SHAP analysis results for different feature categories. CAD factors consistently had the greatest influence on both mortality and MACE predictions across all timepoints and modelling cohorts. This influence tended to decrease when predicting long-term outcomes, with the greatest decrease observed in the Cohort 1 models between 90-day (38.2%) and 5-year mortality (15.8%). Features derived from angiography data and patient diagnostic histories were also found to be particularly influential, where angiography data were more important in predicting mortality outcomes and diagnostic history had a greater influence on MACE predictions. Lifestyle factors and demographic features were the least important features for all models, accounting for less than 10% of feature contributions in all models. Comorbidities were more important for predicting MACE than mortality. Although the feature importance values followed the same general trends for each outcome, the relative contributions of each feature category varied between cohorts. While the contributions of the CAD factors and angiography data fluctuated the most, these changes did not appear consistent across all timepoints for either MACE or mortality.

**Figure 3.**
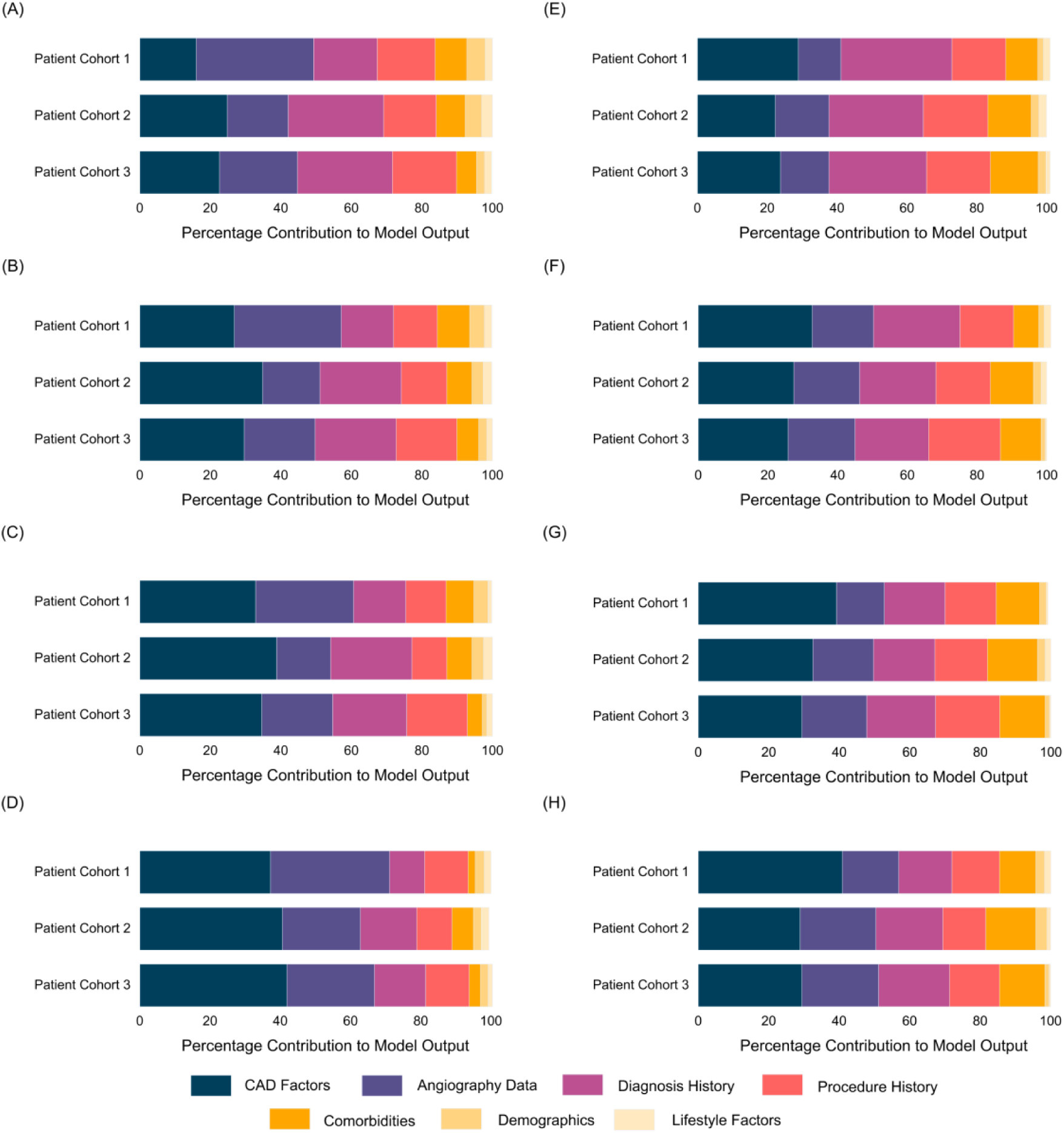
Percentage contributions of major feature categories to absolute mean SHAP feature importance for the selected (A) 5-year mortality, (B) 3-year mortality, (C) 1-year mortality, (D) 90-day mortality, (E) 5-year MACE, (F) 3-year MACE, (G) 1-year MACE, and (E) 90-day MACE prediction models.

Table 3 reports the algorithmic bias assessment results for the best models. While most disparity scores were between 0.8 and 1.25 and hence considered fair, most models’ FPRs were biased and substantially higher in female than male patients. The short-term MACE prediction models (i.e., 90-day and 1-year) tended to be fairer than others.

**Table 3.** Algorithmic bias assessment results, with respect to patient sex, for the best models shown in Table 2. Male was used as the reference group. Asterisks indicate failure to pass the fairness test at a threshold of 0.8 (values between 0.8 and 1.25 are considered fair).

| Test Cohort | FPR Disparity | TPR Disparity | PPV Disparity |
| --- | --- | --- | --- |
| **5-Year All-Cause Mortality** |  |  |  |
| 1 | 1.42* | 1.20 | 0.85 |
| 2 | 1.92* | 1.69* | 1.17 |
| 3 | 1.27* | 1.10 | 1.08 |
| **3-Year All-Cause Mortality** |  |  |  |
| 1 | 1.38* | 1.41* | 1.00 |
| 2 | 1.62* | 1.10 | 1.22 |
| 3 | 1.25 | 0.90 | 1.07 |
| **1-Year All-Cause Mortality** |  |  |  |
| 1 | 1.31* | 1.23 | 1.14 |
| 2 | 1.68* | 1.16 | 1.22 |
| 3 | 1.33* | 1.12 | 1.28* |
| **90-Day All-Cause Mortality** |  |  |  |
| 1 | 1.56* | 1.23 | 1.77* |
| 2 | 1.47* | 1.21 | 1.09 |
| 3 | 1.28* | 1.76* | 1.36* |
| **5-Year MACE** |  |  |  |
| 1 | 1.51* | 1.15 | 0.99 |
| 2 | 1.32* | 1.21 | 1.01 |
| 3 | 1.82* | 1.28* | 0.99 |
| **3-Year MACE** |  |  |  |
| 1 | 1.12 | 1.05 | 1.05 |
| 2 | 1.67* | 1.42* | 1.03 |
| 3 | 1.37* | 1.11 | 1.01 |
| **1-Year MACE** |  |  |  |
| 1 | 1.08 | 0.98 | 1.05 |
| 2 | 1.54* | 1.32* | 1.05 |
| 3 | 1.23 | 1.07 | 1.03 |
| **90-Day MACE** |  |  |  |
| 1 | 1.18 | 1.07 | 1.23 |
| 2 | 1.26* | 1.15 | 1.12 |
| 3 | 1.15 | 1.19 | 1.23 |

FPR—false positive rate; MACE—major adverse cardiovascular events; PPV—positive predictive value; TPR—true positive rate.

## Discussion

### Principal Findings

Using a comprehensive multi-center data set spanning 10 years and over 44,000 catheterizations extracted from the APPROACH Registry and linked administrative health databases, we successfully developed a set of ML models capable of predicting a wide range of short-term and long-term all-cause mortality and MACE outcomes. External validation showed the models’ state-of-the-art prediction performance, very good calibration, and reasonable model fairness across male and female patients.

In general, MACE was more difficult to predict accurately than mortality, presumably due to its composite nature that included 5 disparate endpoints. Also, low event rates, except 5-year MACE, made it challenging for the models to perform well in terms of TPR (a.k.a. sensitivity) and PPV, even with SMOTE, particularly in the cases of 90-day and 1-year mortality where the event rates were less than 3% and 6%, respectively. Some performance variations across sites were also observed even though all three sites are under the same provincial health authority, implying clinical practice variations exist. For both mortality and MACE, CAD factors, angiography results, and patient history had the most influence on predictions overall.

For most outcome-test cohort combinations, TabPFN outperformed the other traditional ML models, leveraging its transformer architecture resembling large language models. The use of TabPFN is one of the present study’s main contributions, as TabPFN was introduced only recently and has not been utilized in health research much. TabPFN was able to achieve state-of-the-art performance out-of-the-box without fine-tuning or hyperparameter tuning. Furthermore, TabPFN can handle data preprocessing, imputation, and class imbalance mitigation natively, making it easy to use. TabPFN’s weaknesses include long inference times as training and test sets increase in size, and substantial memory and compute resource requirements.

The secondary analysis showed that prediction performance generally did not degrade meaningfully when angiography features were excluded. However, in general, the best prediction performances were achieved when angiography features were included. Hence, while it would be preferred to use angiography data to maximize prediction performance, if it is challenging to access them in real-time at the point of care, prediction performance without them is still close to optimal performance.

Algorithmic bias analysis was a strength in our study since such analysis is still rare in health ML studies. Our results increase the trust in our models by demonstrating that they are mostly fair across male and female patient groups. However, most models were deemed unfair in terms of FPR disparity, with greater FPRs in females than males meaning females are more likely than males to be incorrectly predicted to result in a MACE or mortality event when the ground truth is negative. This was presumably caused by the fact that approximately three quarters of the patients in our data set were male. This model bias may lead to unnecessarily aggressive, proactive treatments for females, which can be viewed as less concerning than undertreatment.

### Clinical Implications

Accurate prediction of important patient outcomes can greatly enhance treatment planning for patients with CAD. In particular, risk probabilities generated by our ML models conditioned on treatment (e.g., what is the probability of 5-year MACE if this patient were to receive PCI?) can enable a direct comparison among the treatment options. While clinical practice guidelines exist to aid coronary revascularization decision-making, they are not meant to be personalized and are sub-optimal for many unique patients. ML models that can ingest and analyze complex patient data are likely to outperform clinicians in outcome prediction at the individual patient level, particularly for long-term outcomes. Improved revascularization decisions supported by ML-based predictions have the potential to enhance patient quality of life, reduce health care costs, and increase clinicians’ confidence and satisfaction via better patient outcomes and prevented adverse events.

### Comparisons with Previous Studies

Our models improve the state-of-the-art in CAD patient outcome prediction in several ways. First, our models were developed using a large multi-center cohort of diverse patients with CAD with very few exclusion criteria, allowing for more robust, well-performing models externally validated with patient data that better represent real-world clinical practice. Most previous studies were based on smaller single-center data with strict inclusion and exclusion criteria, jeopardizing their real-world applicability. Second, our models predicted a wide range of short-and long-term patient outcomes, whereas previous studies tended to focus on only one specific outcome. Third, our models are presumably more personalized given that they utilized 200 features selected from over 12,000. Even the ML studies discussed below were based on total feature pools of fewer than 100. Fourth, the fairness of our models was established. To the best of our knowledge, algorithmic bias analysis has never been conducted in outcome prediction research in patients with CAD. Fifth, while previous studies only utilized traditional statistical and ML models, most of our best models were modern, transformer-based TabPFN models.

Our models (AUROC ranged 0.797-0.845 and 0.694-0.753 for mortality and MACE, respectively) substantially outperformed traditional risk scoring systems for patients with CAD. For example, the SYNTAX score has shown poor discriminative performance when used to predict long-term mortality and MACE, with reported AUROCs generally falling between 0.50 and 0.65.^16–19^ Extensions of the SYNTAX score incorporating additional clinical variables, such as the clinical SYNTAX, SYNTAX II, and SYNTAX II 2020 scores, have shown an increased ability to predict long-term MACE and mortality outcomes, with reported AUROCs ranging from 0.60 to 0.80.^2,16,18–21^ However, those studies reporting AUROCs near the upper end of this range generally carried out validation in a small cohort of homogeneous patients unlike this present study.^16,21^ Moreover, the extreme variation in reported AUROC values suggests that conventional risk scores may lack generalizability. This might be explained by the fact that these algorithms were mostly developed in patient cohorts derived from clinical trials with strict inclusion and exclusion criteria.^16,17,21^

Outside of SYNTAX-based scores, several risk scores considering only clinical variables, such as the Age, Creatinine, and Ejection Fraction (ACEF) score, EuroSCORE, and Global Registry of Acute Coronary Events (GRACE) score, have also been shown to be useful in predicting outcomes in patients with CAD, with reported AUROCs ranging from 0.69 to 0.80 for mortality and 0.52 to 0.75 for MACE.^17,22–26^ However, these scores may have limited practicality in individualized decision-making for patients with complex CAD as they either 1) were developed as general cardiac surgery risk scores not specific to patients with obstructive CAD and/or 2) consider post-operative variables that cannot be determined during pre-procedural treatment planning.^25–27^

When compared to other ML-based risk prediction studies in the literature, our models performed either better or similarly. For example, a random forest model trained to predict 4-year mortality in patients with three-vessel CAD was found to perform well with an AUROC of 0.81 during internal validation.^28^ Another study describing an XGBoost model targeting 6-month MACE in patients undergoing coronary revascularization reported an AUROC of 0.86 and an F1 score of 0.77.^3^ Kwon et al.^4^ reported an impressive AUROC of 0.99 and an AUPRC of 0.8, but these results were limited to 30-day mortality, only patients receiving either PCI or CABG, and internal validation. Ninomiya et al.^5^ achieved an AUROC of 0.77 for 5-year mortality which is below our model performance. Lastly, Bertsimas et al.^6^ aimed to predict 10-year 2-point MACE (stroke and myocardial infarction) using a single-center data set focused on patients with general CAD risk and achieved an AUROC of 0.815. The generalizability of all these ML studies may again be limited by their small sample sizes (the largest was 21,460 patients^6^ and most other studies used data sets one order of magnitude smaller), single-center nature, absence of external validation, and strict patient exclusion criteria (e.g., only three-vessel disease^28^).

### Limitations and Future Work

This study has several limitations. First, although a multi-center data set was leveraged, all three sites were from one Canadian province. Prediction performance validation outside of Alberta and Canada is still warranted. Second, the MACE definition used in this study is one of many that have been employed in the literature.^29^ Other MACE definitions should be explored to identify those that can be predicted more accurately. Third, while comprehensive features were used, they were all tabular and angiogram images were not utilized. Multi-modal patient outcome prediction using both angiograms and tabular clinical data may be a promising avenue. Fourth, the scope of the study only included algorithmic bias assessment and correction of any identified bias was excluded. Future work may focus on improving the FPR disparity of our models by up-sampling female patient cases and/or down-sampling male patient cases, although it is often impossible to satisfy all fairness conditions.^30^

### Conclusions

We developed state-of-the-art ML models for patients with CAD that can predict both short- and long-term MACE and all-cause mortality. The discriminability, calibration, and fairness of these models were externally validated using a large multi-center data set. The extensive feature set ensures the models are personalized. Our models have potential to improve coronary revascularization decision-making for diverse patients with CAD by bringing salient prognostic information to the point of care.

## Data and Source Code Sharing

The patient data used in this study contains real patient information and cannot be shared without permission from the data custodians, Alberta Health Services and Alberta Health. The source code used in this study cannot be shared due to copyright protection.

## Author Contributions

EB developed and validated the ML models, produced all results, and partially wrote the manuscript. BH, RW, BT, and CJM provided clinical input. BL and DAS extracted and linked the patient data. CLFS and AP provided technical input. JL conceived and designed the study, partially wrote the manuscript, provided resources, and oversaw the project. All authors critically revised the manuscript.

## Data Availability

The patient data used in this study contains real patient information and cannot be shared without permission from the data custodians, Alberta Health Services and Alberta Health.

## References

1. Gaudino M, Andreotti F, Kimura T. Current concepts in coronary artery revascularisation. The Lancet. 2023;401:1611–1628.

2. Takahashi K, Serruys PW, Fuster V, et al. Redevelopment and validation of the SYNTAX score II to individualise decision making between percutaneous and surgical revascularisation in patients with complex coronary artery disease: secondary analysis of the multicentre randomised controlled SYNTAXES trial with external cohort validation. The Lancet. 2020;396:1399–1412.

3. Wang J, Wang S, Zhu MX, Yang T, Yin Q, Hou Y. Risk Prediction of Major Adverse Cardiovascular Events Occurrence Within 6 Months After Coronary Revascularization: Machine Learning Study. JMIR Medical Informatics. 2022;10:e33395.

4. Kwon O, Na W, Kang H, et al. Electronic Medical Record–Based Machine Learning Approach to Predict the Risk of 30-Day Adverse Cardiac Events After Invasive Coronary Treatment: Machine Learning Model Development and Validation. JMIR Medical Informatics. 2022;10:e26801.

5. Ninomiya K, Kageyama S, Shiomi H, et al. Can Machine Learning Aid the Selection of Percutaneous vs Surgical Revascularization? Journal of the American College of Cardiology. 2023;82:2113–2124.

6. Bertsimas D, Orfanoudaki A, Weiner RB. Personalized treatment for coronary artery disease patients: a machine learning approach. Health Care Manag Sci. 2020;23:482–506.

7. Ghali WA, Knudtson ML. Overview of the Alberta Provincial Project for Outcome Assessment in Coronary Heart Disease. On behalf of the APPROACH investigators. Can J Cardiol. 2000;16:1225–1230.

8. Anon. Discharge Abstract Database (DAD) metadata | CIHI Accessed April 11, 2025. https://www.cihi.ca/en/discharge-abstract-database-dad-metadata.

9. Anon. National Ambulatory Care Reporting System (NACRS) metadata | CIHI Accessed April 11, 2025. https://www.cihi.ca/en/national-ambulatory-care-reporting-system-nacrs-metadata.

10. Labrecque Langlais É, Corbin D, Tastet O, et al. Evaluation of stenoses using AI video models applied to coronary angiography. npj Digit Med. 2024;7:1–13.

11. Avram R, Olgin JE, Ahmed Z, et al. CathAI: fully automated coronary angiography interpretation and stenosis estimation. npj Digit Med. 2023;6:1–12.

12. Hollmann N, Müller S, Purucker L, et al. Accurate predictions on small data with a tabular foundation model. Nature. 2025;637:319–326.

13. Chawla NV, Bowyer KW, Hall LO, Kegelmeyer WP. SMOTE: Synthetic Minority Over-sampling Technique. Journal of Artificial Intelligence Research. 2002;16:321–357.

14. Lundberg SM, Lee S-I. A Unified Approach to Interpreting Model Predictions. In: Advances in Neural Information Processing Systems. Vol 30. Curran Associates, Inc., 2017.

15. Saleiro P, Kuester B, Hinkson L, et al. Aequitas: A Bias and Fairness Audit Toolkit. 2019.

16. Farooq V, Van Klaveren D, Steyerberg EW, et al. Anatomical and clinical characteristics to guide decision making between coronary artery bypass surgery and percutaneous coronary intervention for individual patients: Development and validation of SYNTAX score II. The Lancet. 2013;381:639–650.

17. Chung W-J, Chen C-Y, Lee F-Y, et al. Validation of Scoring Systems That Predict Outcomes in Patients With Coronary Artery Disease Undergoing Coronary Artery Bypass Grafting Surgery. Medicine. 2015;94:e927.

18. Farooq V, Vergouwe Y, Räber L, et al. Combined anatomical and clinical factors for the long-term risk stratification of patients undergoing percutaneous coronary intervention: the Logistic Clinical SYNTAX score. European Heart Journal. 2012;33:3098–3104.

19. Garg S, Sarno G, Garcia-Garcia HM, et al. A New Tool for the Risk Stratification of Patients With Complex Coronary Artery Disease. Circulation: Cardiovascular Interventions. 2010;3:317–326.

20. Nam CW, Mangiacapra F, Entjes R, et al. Functional SYNTAX score for risk assessment in multivessel coronary artery disease. Journal of the American College of Cardiology. 2011;58:1211–1218.

21. Hara H, Kogame N, Takahashi K, et al. Usefulness of the updated logistic clinical SYNTAX score after percutaneous coronary intervention in patients with prior coronary artery bypass graft surgery: Insights from the GLOBAL LEADERS trial. Catheterization and Cardiovascular Interventions. 2020;96:E516–E526.

22. Chichareon P, Modolo R, van Klaveren D, et al. Predictive ability of ACEF and ACEF II score in patients undergoing percutaneous coronary intervention in the GLOBAL LEADERS study. International Journal of Cardiology. 2019;286:43–50.

23. Li Y, Li C, Feng D, et al. Predictive value of ACEF II score in patients with multi-vessel coronary artery disease undergoing one-stop hybrid coronary revascularization. BMC Cardiovascular Disorders. 2021;21:489.

24. Wu S, Qiu Z, Lu Y, et al. Predictive value of ACEF II score for adverse prognosis in patients with coronary heart disease after percutaneous coronary intervention. Postgraduate Medical Journal. 2023;99:605–612.

25. Tang EW, Wong C-K, Herbison P. Global Registry of Acute Coronary Events (GRACE) hospital discharge risk score accurately predicts long-term mortality post acute coronary syndrome. American Heart Journal. 2007;153:29–35.

26. Zhao X, Li J, Xian Y, et al. Prognostic value of the GRACE discharge score for predicting the mortality of patients with stable coronary artery disease who underwent percutaneous coronary intervention. Catheterization and Cardiovascular Interventions. 2020;95:550–557.

27. Ranucci M, Castelvecchio S, Menicanti L, Frigiola A, Pelissero G. Risk of Assessing Mortality Risk in Elective Cardiac Operations. Circulation. 2009;119:3053–3061.

28. Feng X, Zhang C, Huang X, et al. Machine learning improves mortality prediction in three-vessel disease. Atherosclerosis. 2023;367:1–7.

29. Bosco E, Hsueh L, McConeghy KW, Gravenstein S, Saade E. Major adverse cardiovascular event definitions used in observational analysis of administrative databases: a systematic review. BMC Med Res Methodol. 2021;21:241.

30. Kleinberg J, Mullainathan S, Raghavan M. Inherent Trade-Offs in the Fair Determination of Risk Scores. 2016.

